# Application of Optimal Control to the Dynamics of COVID-19 Disease in South Africa

**DOI:** 10.1101/2020.08.10.20172049

**Authors:** S. P. Gatyeni, C.W. Chukwu, F. Chirove, Fatmawati, F. Nyabadza

**Affiliations:** Mathematics and Applied Mathematics Department, University of Johannesburg, Auckland Park Kingsway Campus, P O Box 524, 2006, Johannesburg, South Africa; Department of Mathematics, Faculty of Science and Technology, Universitas Airlangga, Surabaya 60115, Indonesia

**Keywords:** COVID-19, South Africa, Optimal control measures, Constant control, Model fitting, Numerical simulations

## Abstract

SARS-CoV-2 (COVID-19) belongs to the beta-coronavirus family, which include: the severe acute respiratory syndrome coronavirus (SARS-CoV) and the Middle East respiratory syndrome coronavirus (MERS-CoV). Since its outbreak in South Africa in March 2020, it has lead to high mortality and thousands of people contracting the virus. Mathematical analysis of a model without controls was done and the basic reproduction number (ℛ_0_) of the COVID-19 for the South African pandemic determined. We introduced permissible controls and formulate an optimal control problem using the Pontraygain Maximum Principle. Our numerical findings suggest that joint implementation of effective mask usage, physical distancing and active screening and testing, are effective measures to curtail the spread of the disease in the human population. The results obtained in this paper are of public health importance in the control and management of the spread for the novel coronavirus, SARS-CoV-2, in South Africa.

## 1. Introduction

The recent pandemic, which is commonly known as novel coronavirus (COVID-19), is a lower respiratory infection which originated from Wuhan China in 2019 and it has spread all over the world [1]. The first case for COVID-19 occurred in early December, 2019 and by January 29, 2020 more than 7000 infected cases had already been reported in China [2]. This virus spreads through coughing, sneezing and most people who fall sick with COVID-19 usually experience mild to moderate symptoms and recover without special treatment [3]. The burden of COVID-19 has been massive worldwide, and governments are finding ways to control the pandemic and protect the health systems of the countries while safeguarding their economies. There is are no antivirus drugs yet to fight the virus and scientists around the world are working on potential treatments and vaccines for the pandemic [4]. The only available mode of minimising the disease burden has been to control its transmission in the population using non-pharmaceutical interventions.

Mathematical models have played a major role in increasing the understanding of the underlying biological and psycho-social processes that control the spread of infectious diseases and how the disease can be managed [5]. Various strategies have to be deployed optimally to manage the spread of COVID-19. It is thus important to consider standard mathematical approaches to determine the optimality of the control strategies. Optimal control is a standard method for solving dynamic optimisation problems, where those problems are defined in continuous time [6]. To formulate an optimal control problem, we need a mathematical model of the system that needs to be controlled, specifications of all the boundary conditions on states and constraints to be satisfied by states and controls [7].

The first case of the COVID-19 disease in South Africa was reported on March 5, 2020 as an imported case from Italy [8]. Due to the increase in new number of cases, the South African government introduced strategies to control the spread of the pandemic such as social distancing to limit close contact among individuals, contact tracing, testing and screening [9]. On March 26, 2020, the South African government enforced a 21-day national lockdown to restrict contact of individuals and to delay the spread of the virus. As of August 3, 2020, the number of infected cases in South Africa for all the provinces was 516 862 out of which 358 037 recovered [10]. When compared to July 3, 2020, the number of cases had increased by 192% and recoveries by 315% in a period of four weeks.

Since the outbreak of the pandemic, researchers have used mathematical and statistical models to predict and model the extent to which the disease can be contained, or controlled. Nyabadza *et al*. [11] considered the impact of social distancing on the spread of COVID-19 in South Africa. Their results showed that an increase in social distancing significantly decreases the disease spread. Masuku *et al*. [12] used a system of differential equations to quantify the early COVID-19 outbreak transmission in South Africa and exploring vaccine efficacy scenarios. Their findings suggested that the COVID-19 outbreak in South Africa had a basic reproductive number of 2.95 and a highly efficacious vaccine would have been required to contain COVID-19 in South Africa. Chandra et al. [13] looked at mathematical models with a social distancing parameter for the early estimation of COVID-19 spread in India. Their results showed that mathematical modelling and simulations on social distancing plays an important role in spread estimation. The effect of social distancing was discussed with different social distancing rates and they found that social distancing can reduce spread of COVID-19. A normalised Susceptible, Exposed, Infected and Recoveries (*SEIR*) mathematical model with optimal control techniques was considered in [14] with the aim of establishing vaccination and treatment plans that reduce the spread of an infectious disease. Their findings revealed that after stopping vaccination and/or treatment, the percentage of the infected people increased. The study in [15] proposed a generalised *SEIR* model of COVID-19 to study the behaviour of the disease transmission under five different control strategies. The results from the numerical simulations reflected that quarantining and better medical treatment of infected individuals reduced the critically infected cases, and the transmission risk.

Emerging evidence and predictions on COVID-19 allude to the fact that the infection may become endemic and may never go away [1]. Any intervention strategies would need to adjust to this new reality. Unlike most of the current models which ignore the vital dynamics, models need to accordingly be adjusted to capture the nature of the pandemic. The current model incorporates the vital dynamics to capture the dynamics of COVID-19 infection using the South African setting in addition to optimizing the control strategies. The objective of this study is to develop dynamics COVID-19 model for the South African setting incorporating current infection prevention efforts that include screening and testing, the use of masks and physical distancing. Optimal control theory is used to quantify the effectiveness of these strategies.

The paper is arranged as follows: in the next section, the relevant model is formulated. Section 3 presents the model properties and analysis without controls. In section 4, the model with optimal control is presented, analysis is done using Pontryagin’s Maximum Principle. Numerical results are presented in section 5 and the last section concludes the paper.

## 2. Methods

### 2.1. Exploratory Data Analysis (EDA) of the COVID-19 in South Africa

We used the open dataset of COVID-19 called Data Repository and Dashboard for South Africa created, maintained, and hosted by the Data Science for Social Impact research group [10]. They made an excellent data repository and dashboard for South Africa to report on daily confirmed cases data. This dataset has daily level information on the number of infected cases, deaths, recoveries, and testing on the confirmed cases of COVID-19. The data was collected from March 03, 2020, when the first case for South Africa was identified. On March 26, 2020, the government implemented a 21-day national lockdown due to increased daily confirmed cases. The restrictions like social distancing, wearing of masks were implemented to control the spread of the disease.

We present data between March 26, 2020, and July 19, 2020. We aim to fit model to COVID-19 data and introduce optimal control measures such as mask usage, physical distancing, active screening, and testing to control the spread of the disease.

### 2.2. Mathematical Model without Control

In this section, we give a brief model description of the transmission dynamics of COVID-19 without control measures. Considering a non-constant population, we describe the model using a set of five compartments of the susceptibles, *S*(*t*), who are recruited at a constant rate Λ.These individuals can get infected [16], and move into the exposed compartment *E*(*t*). Individuals in this compartment do not show symptoms and cannot transmit the infection. After a period of 5-6 between 5 to 14 days, an exposed individual progresses to become symptomatic and infectious [17]. We assume that the exposed individuals become symptomatic at a rate *γ*. Of those that become symptomatic, a proportion *ρ*, is assumed to be undiagnosed due to system capacity failure and the non-development of severe disease and move to the compartment *I*_*u*_(*t*), while the remainder is diagnosed and move to the compartment *I*_*d*_(*t*). We assume that the individuals in *I*_*u*_(*t*) undergo active screening and testing and when confirmed positive, they move to the *I*_*d*_(*t*) class at a rate *σ*. The diagnosed and undiagnosed infectious humans recover at rates *ω*_1_ and *ω*_2_ respectively and move into the recovered compartment *R*(*t*). Individuals in all compartments are assumed to have a natural mortality rate *µ*. The diagnosed and undiagnosed individuals have additional disease induced death rates *δ*_1_ and *δ*_2_ respectively. The force of infection is defined by

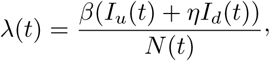

where *β* is the infection rate, and *η* < 1 is the modification parameter used to compare the infectiousness of individuals in *I*_*u*_(*t*) relative to those *I*_*d*_(*t*). Here we assume that the *I*_*d*_(*t*) class infectivity is less that that for the *I*_*u*_(*t*) since individuals who know their status are more likely to limit contact than those who do not know. The total human population *N* (*t*) for the COVID-19 transmission dynamics at any time *t* is given by

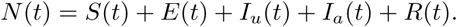

The flow diagram of individuals between compartments is shown in Figure 1. Considering the model description, assumptions, model parameters and Figure 1 we have the following system of ordinary differential equations:

**Figure 1:**
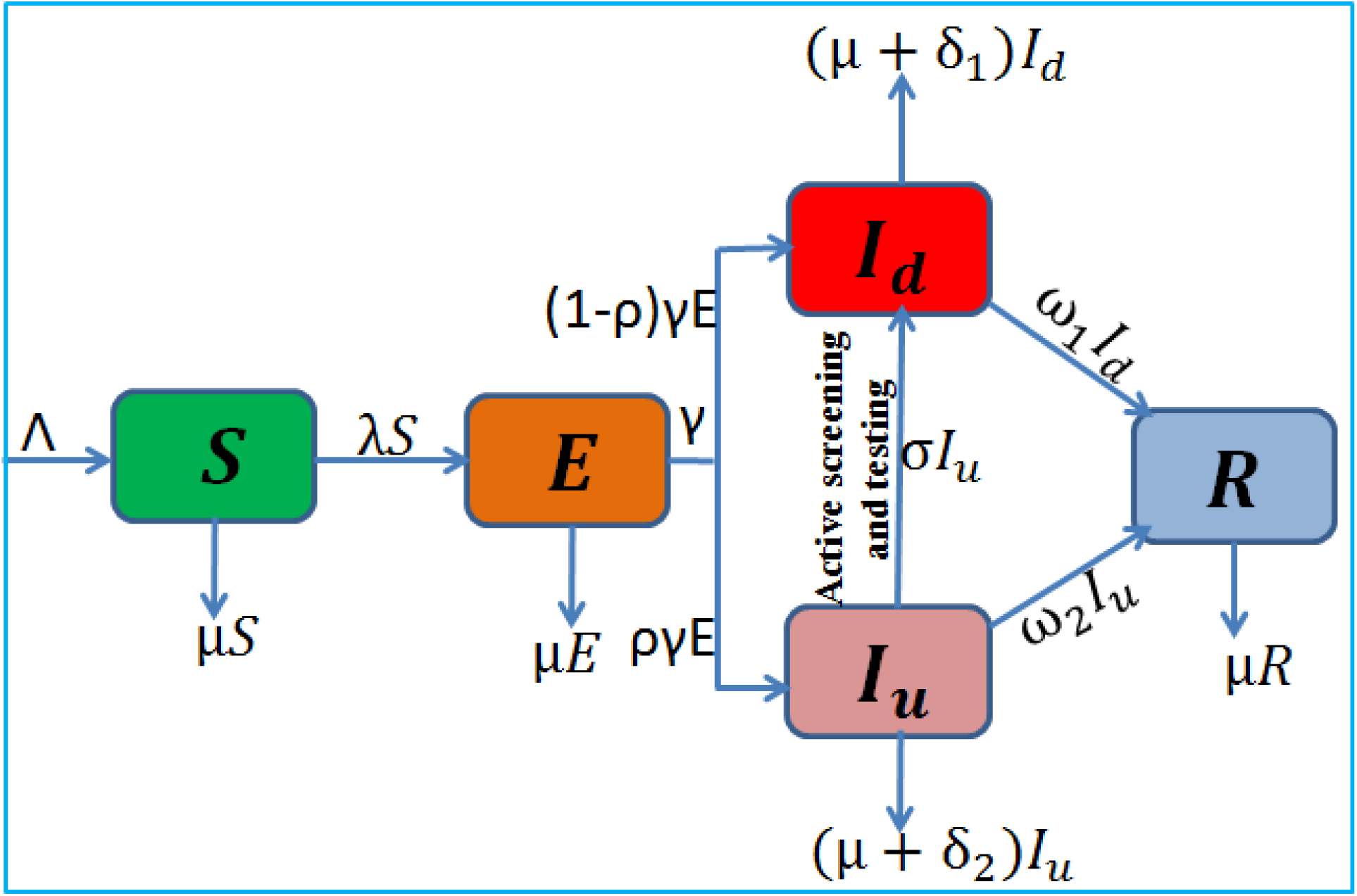
Model flow chart depicting the dynamics of COVID-19 transmissions.

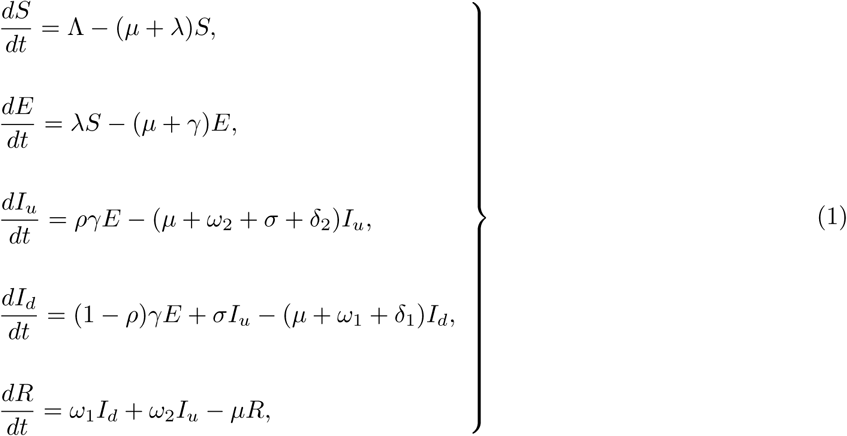

subject to the initial conditions

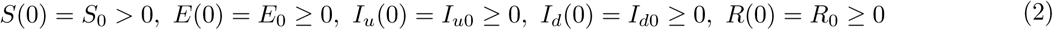

Since the recovered humans do not contribute to COVID-19 disease transmission, we can eliminate *R* from the system using the relationship

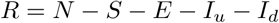

and still adequately represent the dynamics of system (1) by

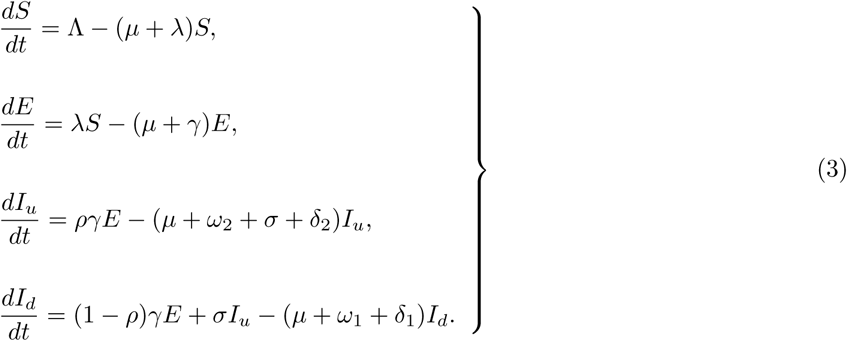

## 3. Mathematical Analysis

### 3.1. Boundedness of solutions

We propose the region Γ in 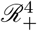 for model equations (3) to be well-posed. We show that these solutions are uniformly bounded. Thus, we have the following theorem

#### Theorem 1

The solutions of model system (3) are contained in the region 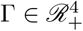, which is given by

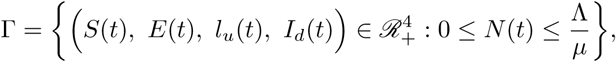

for the initial conditions (2) and attracting in Γ.

*Proof*. The change in total human population of equation (3) is given by

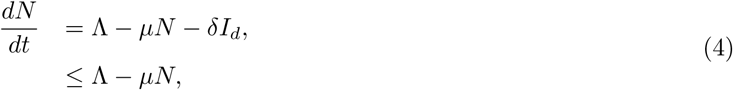

by the positivity of *I*. The solution of (4) yields

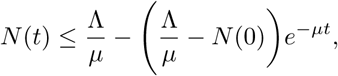

where *N* (0) is a constant representing the the initial condition. Thus, 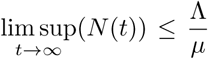. This result implies that the model describing the human population is epidemiologically well-posed, and the solutions remain in the region Γ. □

### 3.2. Positivity of Solutions

Here, we show that the COVID-19 disease model (3) solutions remain positive in the positive orthant given any non negative initial conditions.

#### Theorem 2

Given the initial conditions *S*(0) > 0, *E*(0) ≥ 0, *I*_*u*_(0) ≥ 0, *I*_*d*_(0) ≥ 0, the solutions (*S*(*t*), *E*(*t*), *l*_*u*_(*t*), *I*_*d*_(*t*)) of the system (3) are all positive for all *t* ≥ 0.

*Proof*. From the first equation of the system (3), we have that

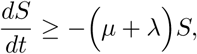

whose solutions gives

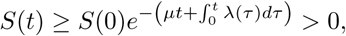

Thus, *S*(*t*) will always be positive for all *t* ≥ 0. Also, from the second equation of the system (3), we have that

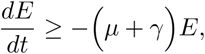

whose solution yields

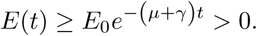

Thus, *E*(*t*) will always be positive for all *t* ≥ 0. Similarly, the solutions of the remainder of the equations in (3) is straight forward and are all positive; that is, *I*_*d*_(*t*) > 0, and *I*_*u*_(*t*) > 0 over the modelling time. □

### 3.3. Equilibrium Points

#### 3.3.1. The Disease Free Equilibrium

Assuming that at the disease free equilibrium (DFE) point there are no infectious individuals. Setting the right hand side to zero, we obtain the DFE denoted by

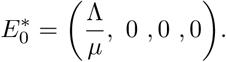

#### 3.3.2. The Reproduction Number

For simplicity, we set *Q*_0_ = (*γ* + *µ*), *Q*_1_ = (*ω*_2_ + *σ* + *δ*_2_ + *µ*), *Q*_2_ = (*ω*_1_ + *δ*_1_ + *µ*) in (3). The basic reproduction number denoted by (ℛ_0_) is the average number of secondary cases generated by an infectious individual who introduced in a wholly susceptible population. We calculate the COVID-19 (ℛ_0_) for model system (3) using the next generation matrix method in [18]. We consider three compartments *E, I*_*u*_ and *I*_*d*_ which contribute to new infections or the secondary cases and construct the column matrices;

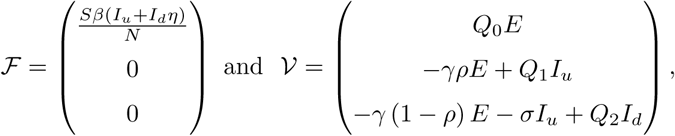

where, ℱ represents the rate of generation of new infections in compartments *E, I*_*u*_ and *I*_*d*_ and 𝒱 represents the transition in and out of the compartments. We define matrices *F* and *V* to the Jacobian matrices for ℱ and 𝒱 respectively evaluated at the DFE. The basic reproduction number is the spectral radius of the product of *F* and *V* ^−1^ i.e, ℛ_0_ = *ρ*(*FV* ^−1^) where *V* ^−1^ in the inverse matrix of the matrix V. Computing the the matrices*F, V* and *V* ^−1^ we obtain;

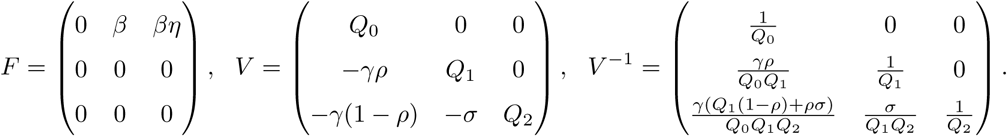

The spectral radius is thus the maximum eigenvalue of the second generation matrix *FV* ^−1^ and is given by

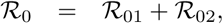

Where

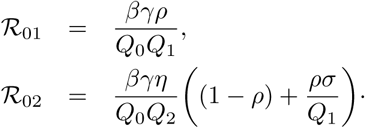

The expression *ℛ*_01_ represents the number of new COVID-19 infections generated by infectious undiagnosed individuals, *I*_*u*_. It contains the product of the transmission rate *β*, the fraction 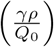 of exposed individuals moving to undiagnosed class *I*_*u*_ and 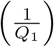 is the average period spend in the *I*_*u*_ class. ℛ_02_ represents the number of new COVID-19 infection cases generated by diagnosed infectious individuals, *I*_*d*_. *βη* is the product of infectious rate due to diagnosed COVID-19 individuals, 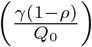 fraction of exposed individuals moving to diagnosed infectious stage and 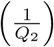 average period spent in the *I*_*d*_ compartment.

It is easy to show that our model satisfy the hypotheses of the Theorem 2 in [18] and hence, the stability of the DFE is ensured and summarised as follows:

##### Theorem 3

The DFE is locally asymptotically stable whenever *ℛ*_0_ < 1 and unstable otherwise.

#### 3.3.3. The Endemic Equilibrium

After some algebraic manipulations, the endemic equilibrium denoted as (EE) is given by

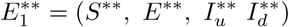

Where

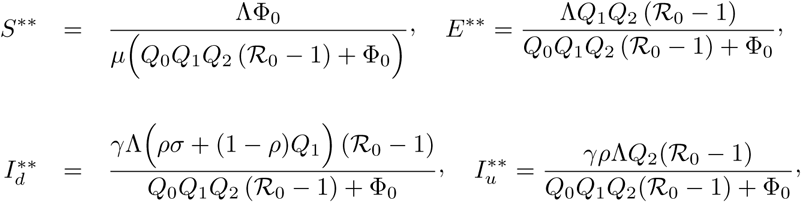

With

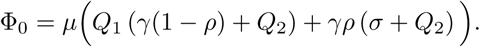

We have the following result on the existence of 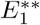.

##### Lemma 1

The EE exist if and only if ℛ_0_ > 1.

### 3.4. Global Stability Analysis of the Equilibria

In this section, we show that the disease-free equilibrium is globally asymptotically stable if *ℛ*_0_ < 1. We also prove that the endemic equilibrium state is globally asymptotically stable if *ℛ*_0_ > 1.

#### Theorem 4

The disease-free equilibrium, *E*_0_ of the model system (3) is globally asymptotically stable if *ℛ*_0_ < 1.

*Proof*. Consider the following Lyapunov function defined by

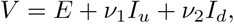

where *ν*_1_ and *ν*_2_ are the positive constants we seek to determine. Finding the time derivative of the function *V* and upon substitution of equations (3) at the DFE we have that

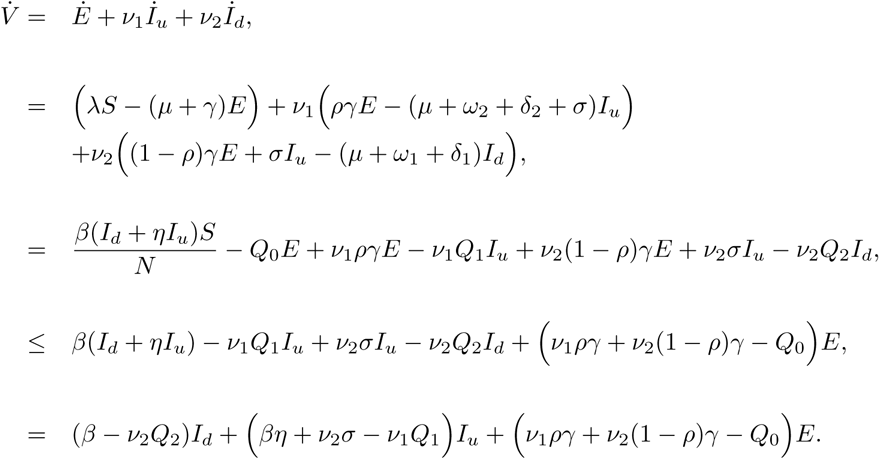

Eliminating *I*_*u*_ and *I*_*d*_ the linear terms, we solve for *ν*_1_ and *ν*_2_ to get

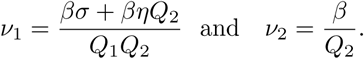

Substituting for *ν*_1_ and *ν*_2_ we obtain

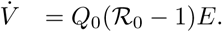

We see that 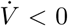 if ℛ_0_ < 1, and 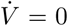 if *I*_*u*_ = *I*_*d*_ = *E* = 0. Therefore, the largest compact invariant set in Γ such that 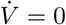 when ℛ_0_ ≤ 1, is the singleton 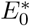. Thus, by LaSalle’s Invariance Principle [19, 20], the steady state 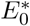 is globally asymptotically stable whenever ℛ_0_ ≤ 1. □

#### Theorem 5

If ℛ_0_ > 1 the endemic equilibrium of the model equation (3), 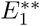 is globally asymptotically stable in Γ.

*Proof*. Given that the endemic equilibrium exists if and only if ℛ_0_ > 1. Consider Lyapunov function

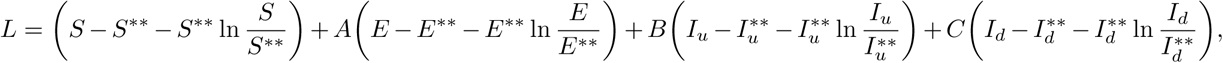

where *A, B* and *C* are constants to be determined. Substituting the expressions for the derivatives of the state variables yields,

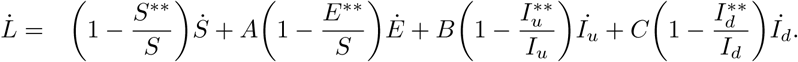

At the steady states we have

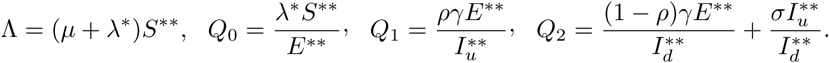

Given that *N* ^∗^ ≤ *N*, and setting 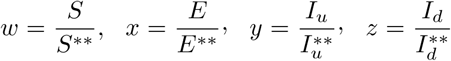, the time derivative of the Lyapunov function is

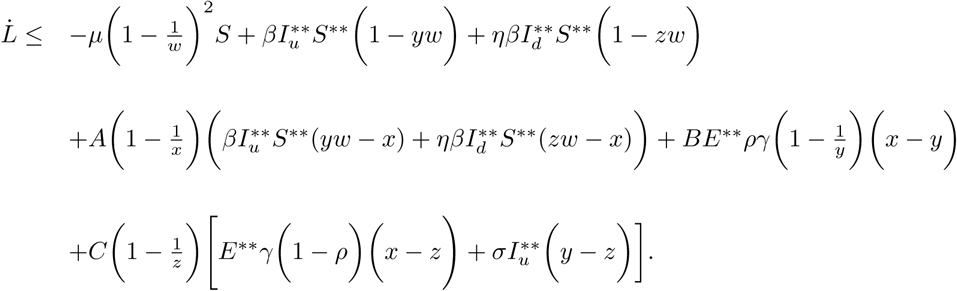

The derivative of *L* thus reduces to

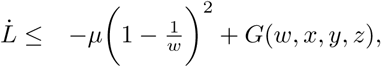

where

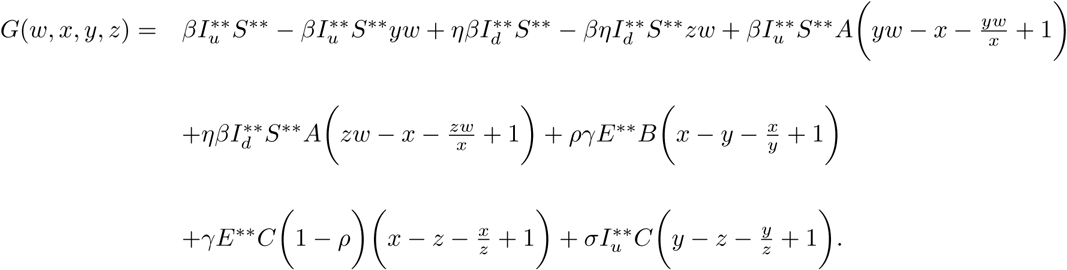

After some algebraic manipulation, we obtain *A* = 1, *C* = *ργE*^∗∗^ and 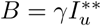 so that

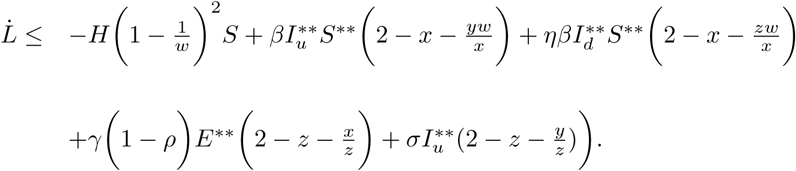

Given that the arithmetical mean is always greater or equal to the geometric mean, the expressions

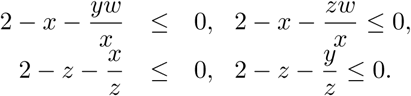

So 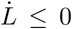 for all *w, y, z* > 0 and 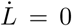 if and only if *S* = *S* ^**^, *E* = *E*^**^, 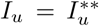 and 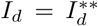. The largest invariant set where 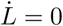 is the singleton of 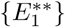. By Lasalle’s invariance principal [20], 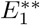 is globally asymptotically stable with respect to the invariant set Γ. □

## 4. Optimal Control Problem

### 4.1. Mathematical Model with Optimal Control

We introduce optimal control measures to the system (3). We formulate an optimal control problem with the following control variables: *u*_1_, the prevention effort of COVID-19 disease such as mask usage, physical distancing, and *u*_2_, active screening and testing. The new force of infection becomes

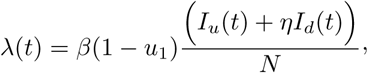

and the control variables to model equation (3) gives rise to

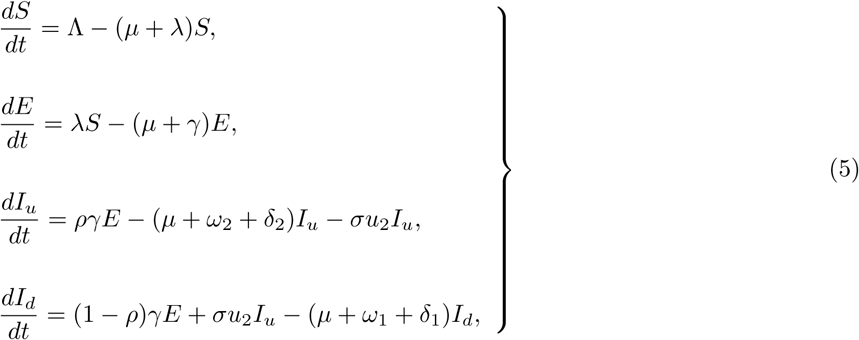

subject to the initial conditions

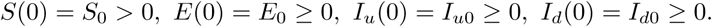

The objective is to reduce the number of undiagnosed humans with the COVID-19 disease. Hence, we formulate a minimization problems with the following objective function

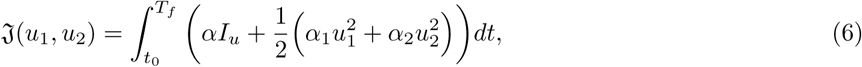

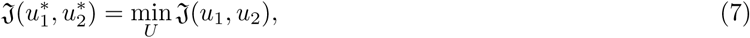

where *t*_0_ is the initial time, *T*_*f*_ is the terminal time, *α* is the weight associated with reducing the undiagnosed infected human class *I*_*u*_, *α*_1_, *α*_2_ are the associated cost weight with the control variables *u*_1_ and *u*_2_ respectively. We define the Hamiltonian function by applying the Pontryagin’s Maximum Principle [21] as follows:

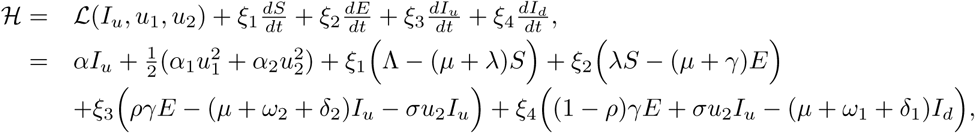

where *ξ*_*i*_, *i* = 1, *…*, 4 are the adjoint variables. We now give the theorem for the existence and uniqueness of the optimal control problem as in [6] as follows;

#### Theorem 6

There exist an optimal control 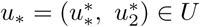 such that

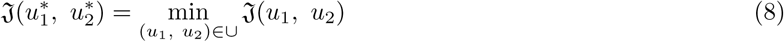

subject to the optimal control model system (5) with initial conditions (1).

### 4.2. Optimality of the System

#### Theorem 7

Given that *S*^*^, *E*^*^, 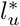 and 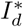 are the solutions of the optimal control model system (5) and (6) associated with the optimal control variables 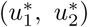. Then, there exist an adjoint system which satisfies

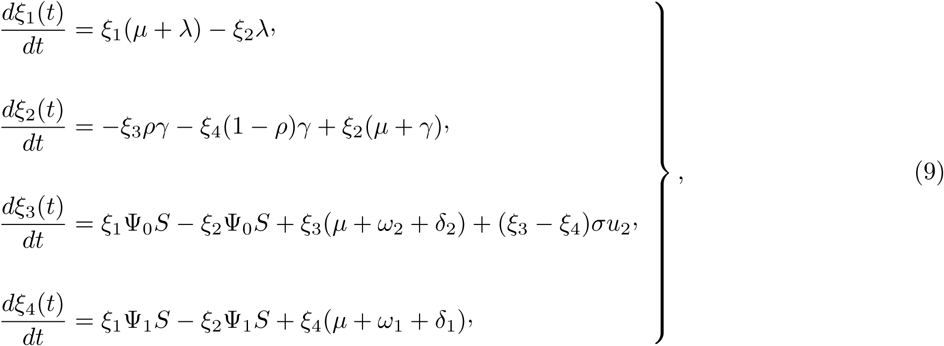

Where

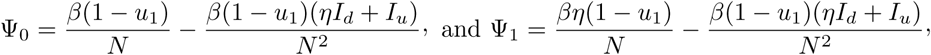

with transversality boundary conditions defined as

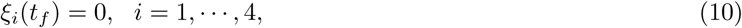

given by

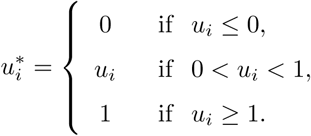

where the permissible control functions 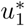, and 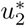, are obtained by setting 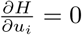, *i* = 1,2. Thus

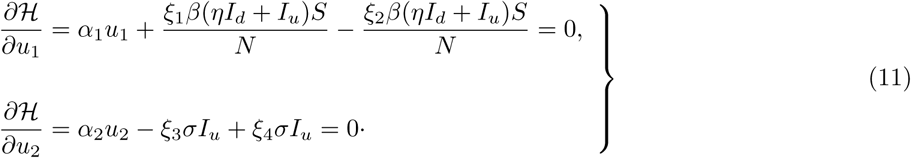

From equation (11), we solve for 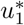 and 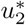 to obtain their respective permissible control, solutions, which are

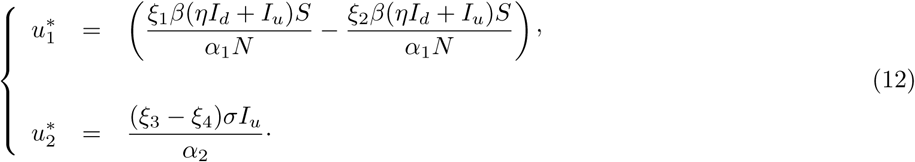

With the upper and lower constraints on the admissible controls 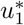, and 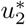, we now have that the optimal controls can be characterised as

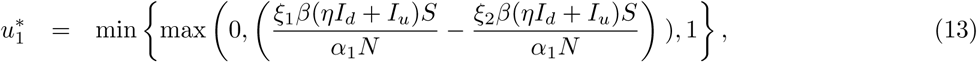

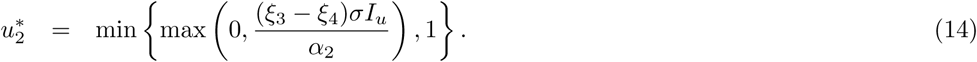

### 4.3. Model Fit with Controls and Parameter Estimation

We use the method of least square curve fitting to fit system (3) to the data. The method is useful in obtaining the parameters values that are then used in the numerical simulations. Figure 2 shows that model (3) fits well to the COVID-19 new daily cases of South Africa as from the 26^*th*^ of March up to the 19^*th*^ of July 2020, with data obtained from [10].

**Figure 2:**
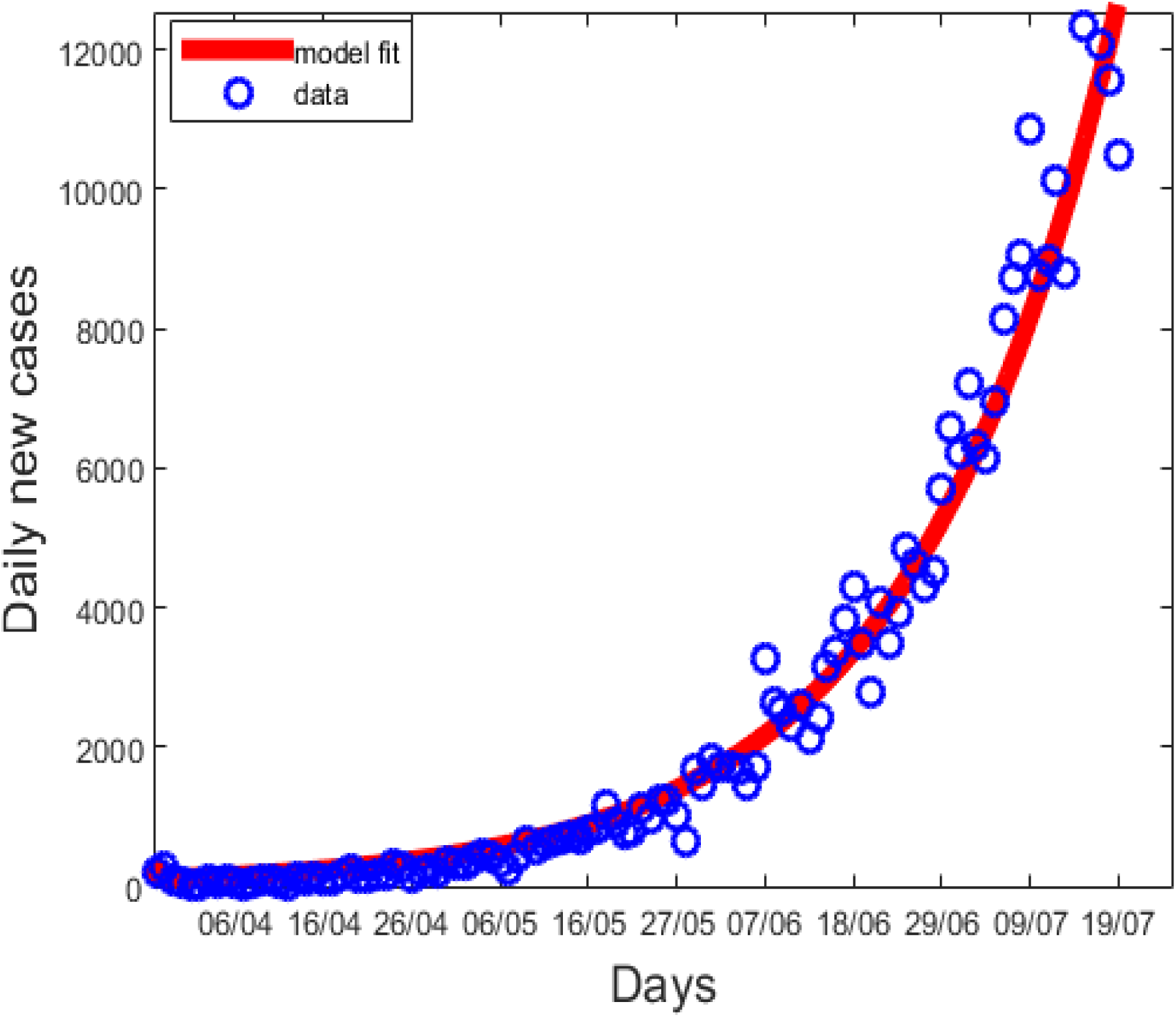
Model fit to data for COVID-19 early growth of new cases in South Africa for the first wave from 26 March to 19 July 2020. Optimal parameter values: Λ = 11244, *µ* = 0.0091, *σ* = 0.2070, *δ*_1_ = 0.01500, *δ*_2_ = 0.0089, *η* = 0.5663, *γ* = 0.5732, *ρ* = 0.9799, *ω*_1_ = 0.1800, *ω*_2_ = 0.0206, *β* = 1.3295, *u*_1_ = 0.7557, *u*_2_ = 0.3728, *ℛ*_0_ = 2.0109.

The curve fitting process generates the following parameters with some of the parameters having been obtained from the cited research.

### 4.4. Sensitivity Analysis

We perform sensitivity analysis of model (3) using the methodology described in [24]. In this paper, the objective of sensitivity analysis is to identify critical parameters driving the COVID-19 pandemic in South Africa. Figure 3(a) shows the effects of parameters *σ, γ, ρ, β, u*_1_ and *u*_2_ on undiagnosed infectious *I*_*u*_ population. *σ* is largely negatively correlated to *I*_*u*_. The increase in the rate of active testing is associated with the decrease of undiagnosed infectious class. *γ* is mostly negative correlated to *I*_*u*_. The increase in the rate at which exposed become infectious is associated with the increase in the undiagnosed class. *ρ* is mostly positive correlated to *I*_*u*_. The increase in proportion of exposed individuals which becomes undiagnosed is associated with the increase in the undiagnosed class. *β* is mostly positive correlated to *I*_*u*_. The increase in the infection rate is associated with the increase in the undiagnosed class. *u*_1_ is negatively correlated to *I*_*u*_. The increase in the mask usage and physical distancing is associated with the decrease in the undiagnosed class. *u*_2_ is negatively correlated to *I*_*u*_. The increase in the active testing and screening is associated with the decrease in the undiagnosed class. Figure 3(b) shows the effects of parameters *σ, γ, ρ, β, u*_1_ and *u*_2_ on diagnosed infectious *I*_*d*_ population. *σ* is largely negatively correlated to *I*_*d*_. The increase in the rate of active testing is associated with the increase of diagnosed infectious class. *γ* is mostly positive correlated to *I*_*d*_. The increase in the rate at which exposed become infectious is associated with the increase in the diagnosed class. *ρ* is mostly positive correlated to *I*_*d*_. The increase in proportion of exposed individuals which becomes undiagnosed is associated with the decrease in the diagnosed class. *β* is mostly positive correlated to *I*_*d*_. The increase in the infection rate is associated with the increase in the diagnosed class. *u*_1_ is negatively correlated to *I*_*d*_. The increase in the mask usage and physical distancing is associated with the decrease in the diagnosed class. *u*_2_ is negatively correlated to *I*_*d*_. The increase in the active testing and screening is associated with the decrease in the diagnosed class.

**Figure 3:**
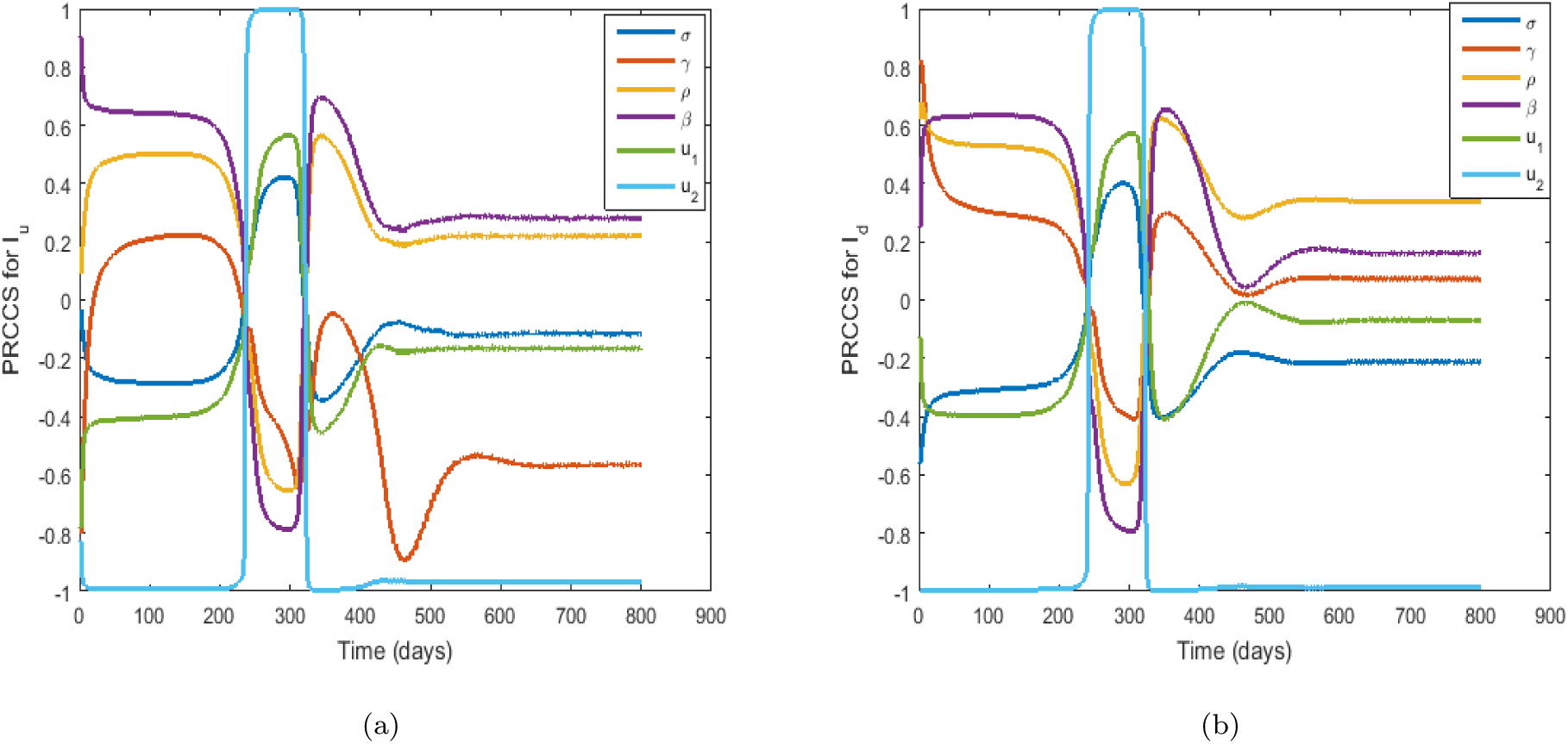
PRCC plots showing the effects of the dynamics of parameter values on COVID-19 daily new cases in South Africa for (a) parameters *β, ρ, σ* and *γ* on *I*_*u*_. (b) parameters *β, ρ, σ* and *γ* on *I*_*d*_.

### 4.5. Effects of Parameters on the ℛ_0_

The effect of parameter on ℛ_0_ were analysed using a contour plot. We chose two significant parameters *γ* and *σ* and give the contour plot as a function of ℛ_0_. Figure 4 shows that an increase in the parameter *σ* results in a decrease in the numerical value of ℛ_0_. This implies that increased screening has the propensity to decrease the spread of COVID-19. It is thus important to increase the rate of screening and testing in order to control the pandemic. On the other hand, the parameter *γ* does not significantly impact ℛ_0_.

**Figure 4:**
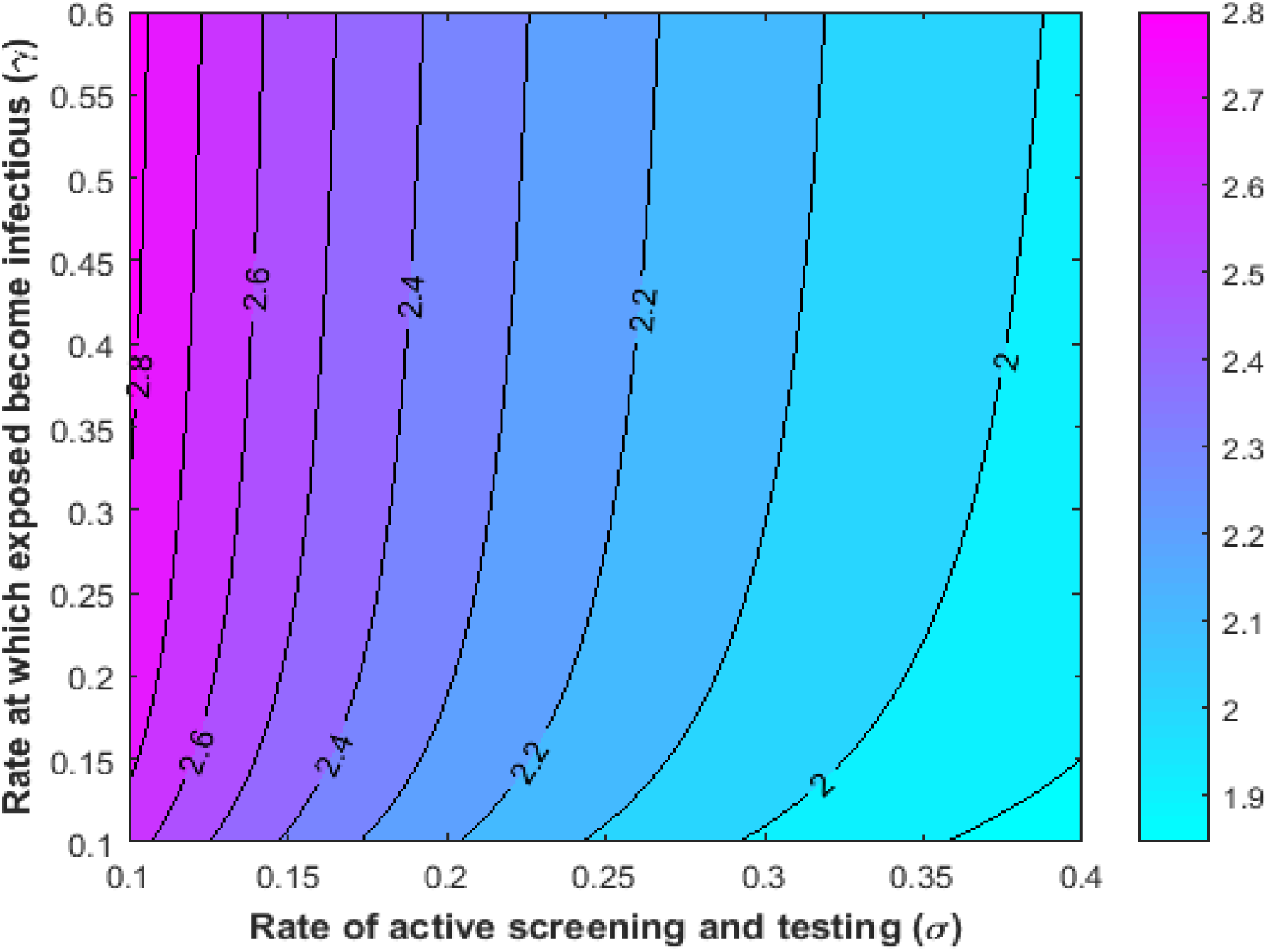
Contour plot of ℛ_0_ as a function of *γ* versus *σ*.

## 5. Numerical Results

The optimal control strategy is found by the iterative method of the fourth order Runge-Kutta iterative scheme. The state equations are initially solved by the forward Runge-Kutta method of the fourth order. Then, by using the backward Runge-Kutta method of the fourth order (backward sweep), we solved the co-state equations with the transversality conditions. The permissible controls are updated by using a convex combination of the previous controls and the value from the characterisations of 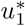 and 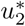. This process is re-iterated and the iteration is ended if the current state, the adjoint and the control values converge sufficiently following [6]. Also, we perform numerical simulations based on the control variables, to investigate the effects of introducing time dependent controls. We consider the combination of the following three different scenarios, enumerated as follows: (1) the use of only one permissible control *u*_1_, (2) the use of only one permissible control *u*_2_, and (3) the use of a combination of permissible controls *u*_1_ and *u*_2_. Using the model parameter values in Table 1, the initial conditions as defined in Section 4.3 and the associated weights from Tables 2 to simulate the model with controls and obtain the following results.

**Table 1:** Parameters of (COVID-19) *SEIR* model for South Africa and their values.

| Symbol | Parameter description | Value [Range] day<sup>-1</sup> | Source |
| --- | --- | --- | --- |
| $\Lambda$ | Recruitment rate | 11244 [10000 - 25000] | Estimated in[11] |
| $\mu$ | Mortality rate | 0.0091 [0.0050 - 0.010] | [22] |
| $\sigma$ | Rate of active testing | 0.2070 [0.4000 - 1.000] | Data fit |
| $\delta_1$ | Diagnosed induced death rate | 0.1500 [0.0000 - 0.100] | [23] |
| $\delta_2$ | Un-diagnosed induced death rate | 0.0089 [0.0000 - 0.100] | Data fit |
| $\eta$ | Modification parameter | 0.5663 [0.0000 - 1.000] | Data fit |
| $\gamma$ | Rate at which exposed become infectious | 0.5732 [0.2000 - 0.600] | Data fit |
| $\rho$ | Proportion of $E$ which becomes undiagnosed | 0.9799 [0.0000 - 1.000] | By definition |
| $\omega_1$ | Recovery rate of diagnosed infected humans | 0.1800 | Data fit |
| $\omega_2$ | Recovery rate of undiagnosed infected humans | 0.0206 [0.0000 - 0.100] | [23] |
| $\beta$ | Infection rate | 1.3295 [1.2100 - 1.500] | Data fit |
| $u_1$ | Mask usage and physical distancing | 0.7557 [0.0000 - 1.000] | Data fit |
| $u_2$ | Active screening and testing | 0.3728 [0.0000 - 1.000] | Data fit |

**Table 2:** Costs associated with optimal control variables used for simulations

| Symbol | Parameter description | Base line Values (day<sup>-1</sup>) | Source |
| --- | --- | --- | --- |
| $\alpha$ | Weight associated with reducing the undiagnosed infected human | 1 | Estimated |
| $\alpha_1$ | Weight on the cost of the prevention | 0.9 | Estimated |
| $\alpha_2$ | Weight on the cost of the active screening and testing | 0.5 | Estimated |

### 5.1. Scenario (1)

This scenario shows the dynamics of COVID-19 using the control variable *u*_1_, which consist of mask usage and physical distancing to optimise the objective function 𝔍, while setting the control variable *u*_2_ to zero. We observed that in Figure 5(a), that maintaining a strict masks usage and physical distancing have significant influence on the peaks of undiagnosed cases of COVID-19. In the absence of active screening and testing, masks and social distancing has the propensity to minimise the spread of COVID-19 disease in South Africa. Figure 5(b), shows the control profile *u*_1_ for this particular first scenario, where *u*_1_ is is given the maximum of approximately 20 days to drop to its lower bound, translating to a maximum effectiveness of masks usage and social distancing.

**Figure 5:**
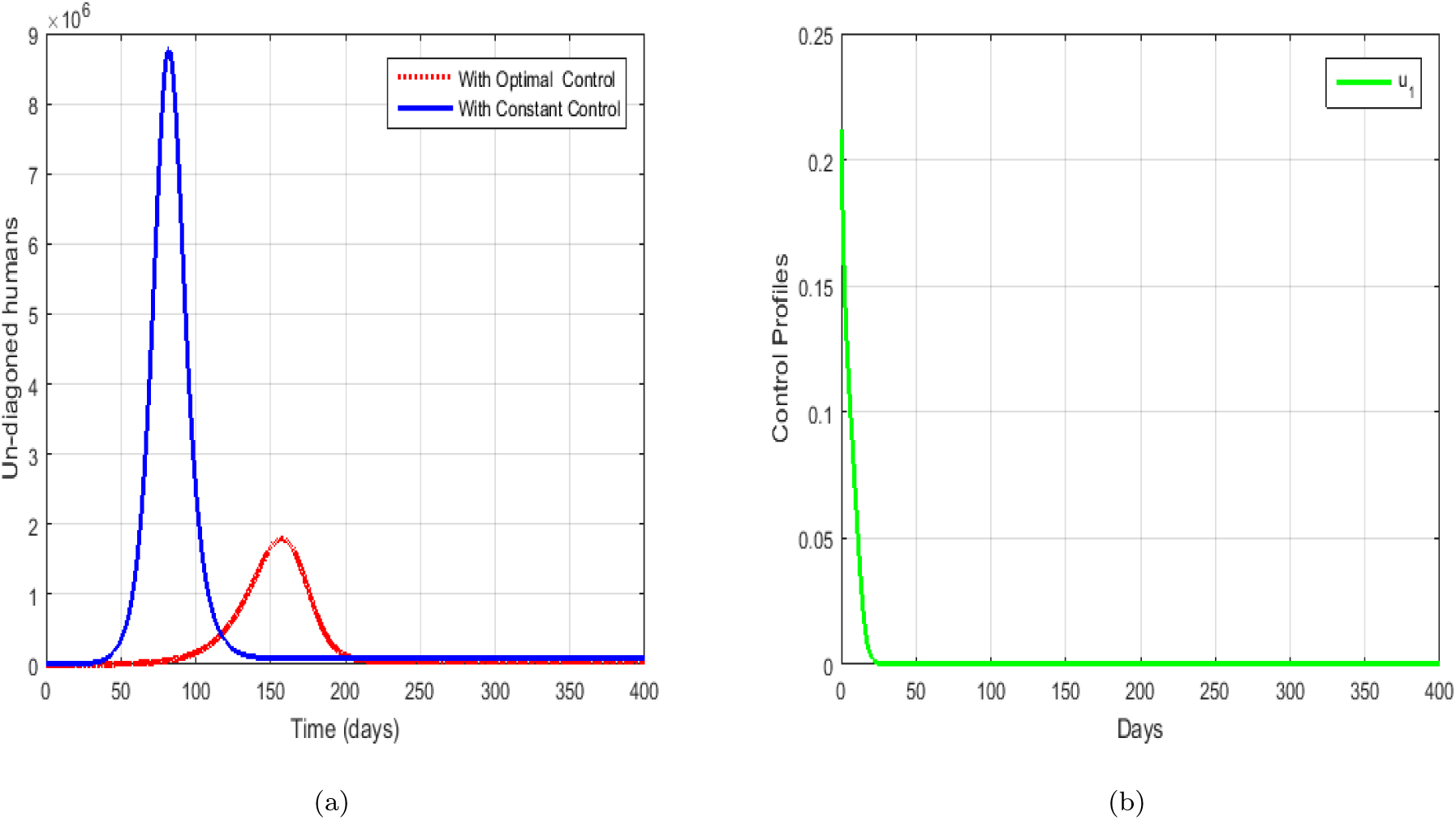
The dynamics on COVID-19 using control 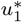 with 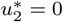 for (a) undiagnosed humans (b) the control profile for 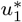.

### 5.2. Scenario (2)

This scenario presents the dynamics of COVID-19 using the control variable for screening and testing *u*_2_ to optimise the objective function 𝔍, while *u*_1_ is set to zero. We observed from Figure 6(a), screening and testing reduces the peak of undiagnoseed infectives. Figure 6(b) shows the control profile for *u*_2_, where *u*_2_ should be allowed to run at its highest level during the entire duration of the pandemic. This means that the scaling of screening and testing should always be done at capacity to control COVID-19.

**Figure 6:**
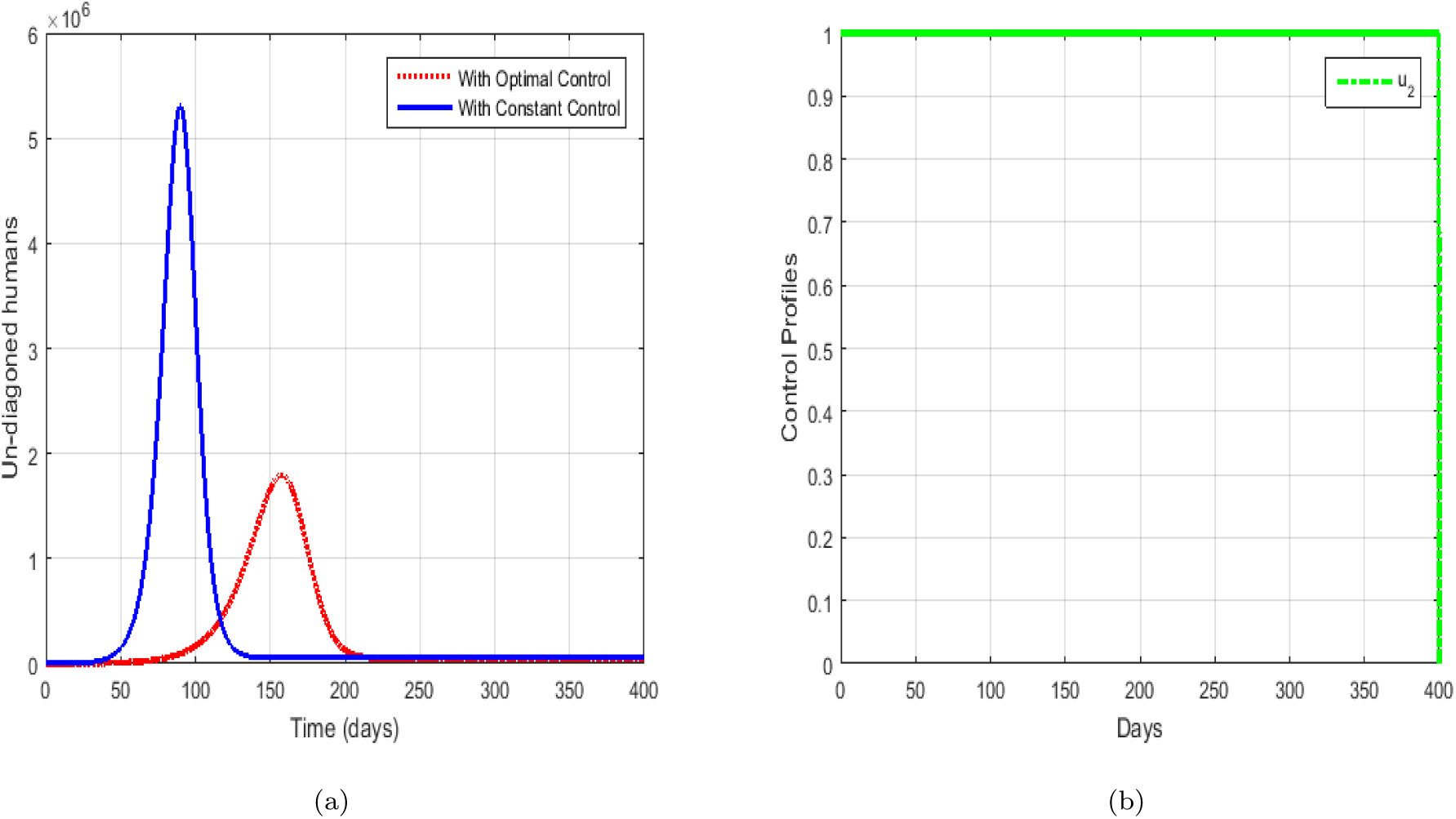
The dynamics on COVID-19 using control 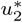, with 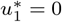 for (a) undiagnosed humans (b) the control profile for 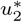.

### 5.3. Scenario (3)

In this scenario, we show both control variables *u*_1_ and *u*_2_ being run concurrently. For these scenarios, we observed in Figure 7(a) that the control strategies of wearing masks, physical distancing and as well screening and testing resulted in a significant decline in the number of undiagnosed COVID-19 cases. Thus, this shows that to reduce the spread of COVID-19, these controls should be used concurrently. Figure 7(b) shows the control profile for both *u*_1_ and *u*_2_.

**Figure 7:**
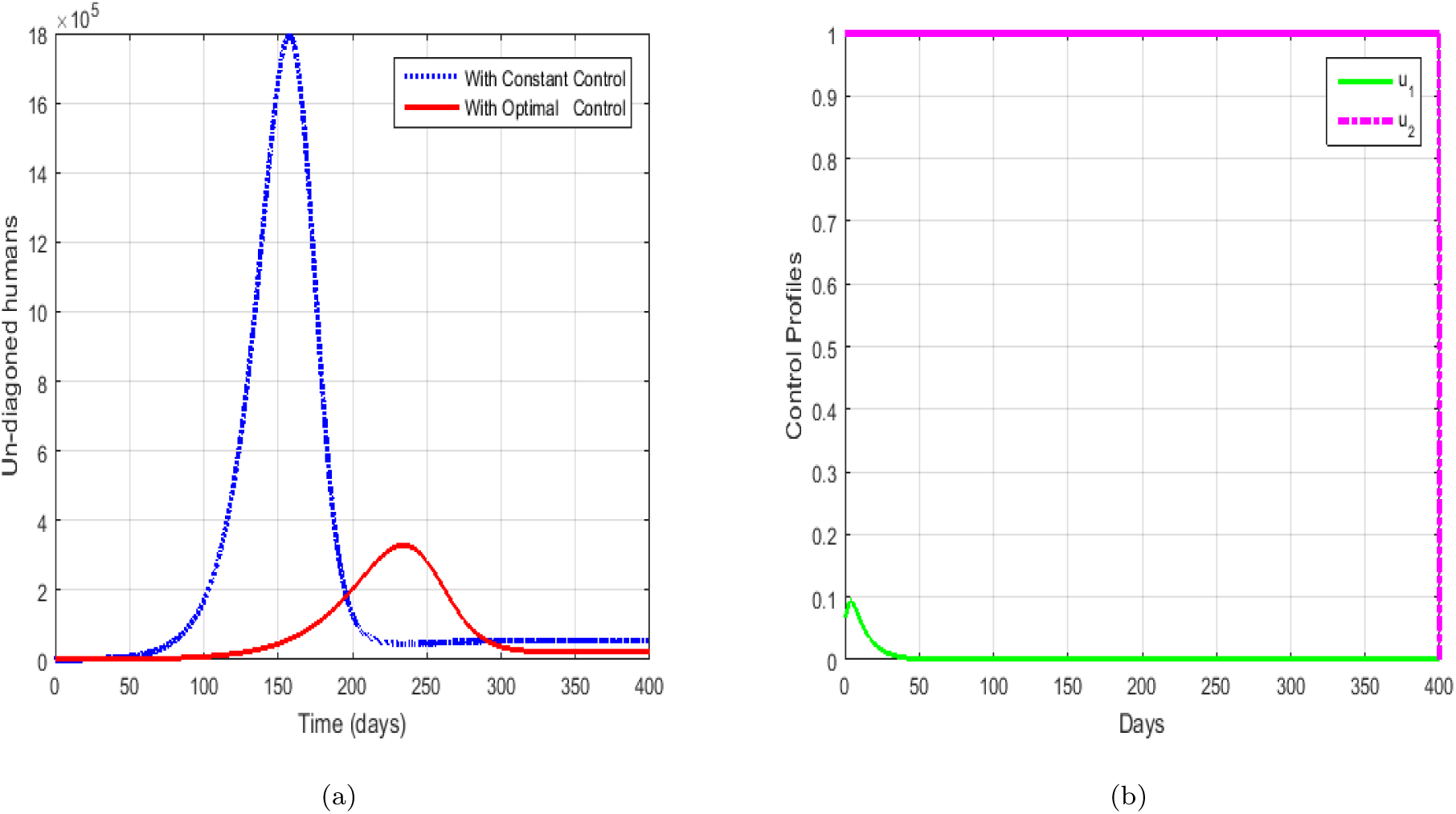
The dynamics on COVID-19 using controls 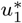 and 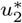 for (a) undiagnosed humans (b) the control profile for 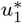. and 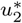.

## 6. Conclusions

In this paper, we used a continuous time model to study the effects of the dynamics of COVID-19 pandemic on the South Africa population. The model takes into account the susceptible, exposed, undetected, detected and recovered humans. We carried out mathematical analysis for the model without controls and showed that the steady steady states are locally asymptotically stable if the basic COVID-19 reproduction number ℛ_0_ is greater than unity and otherwise unstable. Further, we performed the sensitivity analysis of the model equations without control and numerical simulations based on the highly sensitive model parameters on ℛ_0_ as shown in Figures 3. On the other hand, we formulated a time dependent optimal control problem by introducing control variables. The model with control system (5) was solved analytical and numerically to predict the possible and necessary controls to contain the spread of the disease. The results on the numerical simulations suggest that the best strategy is to wear masks and physical distancing *u*_1_, followed by control of testing and screening *u*_2_ to reduce COVID-19 infection and these strategies will be more effective in controlling the rate at which the uninfected humans contract the disease.

While many research papers on COVID-19 have been published to date, the work presented here focuses on the dynamics of the infection in the presence to time dependent optimal controls. Given that new scientific evidence on the virus continues to emerge, the model presented here is certainly neither the ultimate dossier not without culpability. The results should then be treated with circumspection. Some of the major challenges, other than the continuous evolution of scientific evidence, are the none inclusion of some of the dynamics of COVID-19 such as the role played by contaminated surfaces, asymptomatic infectives and the age dependant disease dynamics.

Aspects of stochasticity in the infection rates, the role of social-economic factors in COVID-19 infection spread and the evolution of government policies can be incorporated in the model to improve its usefulness. Also, in [17], it is documented that exposed individuals in compartment *E* can be contagious. Therefore, the assumption that individuals in compartment *E* can not transmit COVID-19 can be relaxed. This means that transmission from individuals in compartment *E* can occur before the onset of symptoms. Such an adjustment requires the inclusion of compartment *E* in the force of infection. Despite these fallibilities, the model still presents some interesting results on the introspections of COVID-19.

## Data Availability

Cited in text

## Data Availability

The data are sourced from public and official government press release [25].

## Conflicts of Interest

No conflict of interest.

## Funding Statement

No funding or grant were utilised for this research work.

## Acknowledgements

The authors would like to thank the Faculty of Science in the University of Johannesburg for material support towards the inception and completion of this project and the University of Universitas Airlangga Indonesia. Author’s will like to appreciate lucrative comments from the anonymous reviewers which can help strengthen the results presented in this research paper. FC, in particular acknowledges the URC grant support towards the completion of the project.

## Notes

### Competing Interest Statement

The authors have declared no competing interest.

### Funding Statement

No fuunding

### Author Declarations

2255

### Summary of Updates

All sections

